# Coxsackievirus A24 variant whole genome sequencing from clinical samples using a three overlapping amplicons strategy

**DOI:** 10.1101/2025.01.16.25320645

**Authors:** John Mwita Morobe, Samuel Odoyo, Arnold W. Lambisia, Edidah Moraa, Charlotte J. Houldcroft, Lynette Isabella Ochola-Oyier, Edward C. Holmes, George Githinji, Charles N. Agoti

## Abstract

We developed a whole genome sequencing assay for coxsackievirus A24 variant (CA24v), a major cause of acute hemorrhagic conjunctivitis, and used it to recover three near complete genomes from the 2024 CA24v outbreak in Kenya. This assay will support studies on CA24v genomic epidemiology and evolution across Africa.

## Background

Coxsackievirus A24 variant (CA24v) is a member of species *Enterovirus coxsackiepol*, genus *Enterovirus*, family *Picornaviridae*, and a leading cause of acute haemorrhagic conjunctivitis (AHC) outbreaks in the tropics, also referred to as “red eye or pink eye” disease ^1–4^. The CA24v genome comprises a single-stranded, positive-sense RNA molecule of approximately 7,400 bp in length and encodes 4 structural proteins (VP4, VP2, VP3, and VP1) and 7 non-structural proteins (2A-2C, and 3A-3D).

As of January 10^th,^ 2024 there were 119 complete or near complete CA24v genomes available in GenBank (>90% coverage) sampled between 1952 and 2024 representing data from 25 countries. This is a small number relative to influenza A (∼165,000), monkeypox virus (∼8,500), SARS-CoV-2 (∼17,000,000) and Ebola virus (∼3,400). The small number of publicly available CA24v genomes is in part explained by limited availability of diagnostic capacity during outbreaks, the self-limiting nature of the AHC condition, and the absence of a simple cost-effective genome sequencing methods ^5,6^. This paucity of CA24b genomic data also limits our understanding of CA24v diversity, evolution and epidemiology ^5^.

Previous efforts to generate CA24v genomes have relied on metagenomic sequencing and primer walking approaches ^6,7^. However, these approaches are relatively expensive, technically demanding, and require significant hands-on time in the laboratory ^8^. An alternative approach is an overlapping amplicon sequencing strategy in which the pathogen genome is amplified as series of tiled fragments which are then sequenced and reassembled ^9^. This strategy has been successfully used on several viral pathogens, including enteroviruses such a Rhinovirus-A15 and A105 ^10^, Enterovirus D68 ^11^, Echovirus 30 ^12^ and Coxsackievirus B5 ^13^. Herein, we present a tiled amplicon approach for CA24v sequencing.

## Methods

We used the *Primal scheme* algorithm with default parameters ^9^ and identified 12 primers (six pairs) that could bind to various positions within the CA24v genome. The input alignment utilized currently available CA24v genomes (>95% coverage) in GenBank. This selection of primers aimed to have pairs that produce an amplicon size of ∼2500 nucleotides. Following laboratory optimisation, six primers that result in three overlapping amplicons were selected (**Table 1**; **Figure 1A**). The resultant amplicons had overlapping regions of 222 nt between amplicon 1 and 2, and 325 nt between amplicon 2 and 3 (**Table 1; Figure 1A**). This set was used to amplify three CA24v positive samples identified in early February 2024 on the Kenyan Coast ^15^ that had a diagnostic cycle threshold (Ct) of 32.29, 38.43, and 37.65 following qPCR.

**Table 1.**
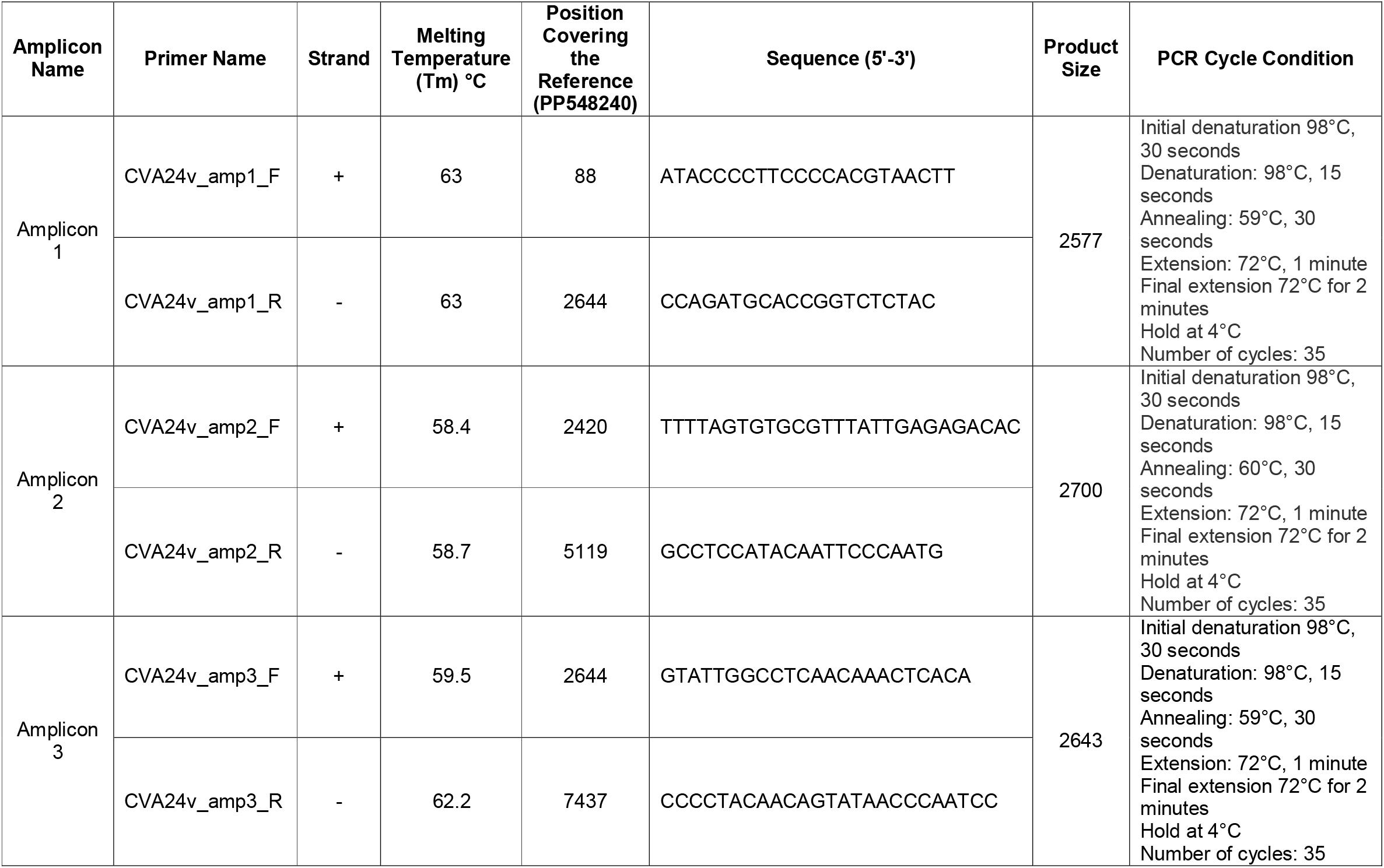
Characteristics of the six primers optimised for CA24 whole genome amplification and the PCR thermocycling conditions.

**Figure 1.**
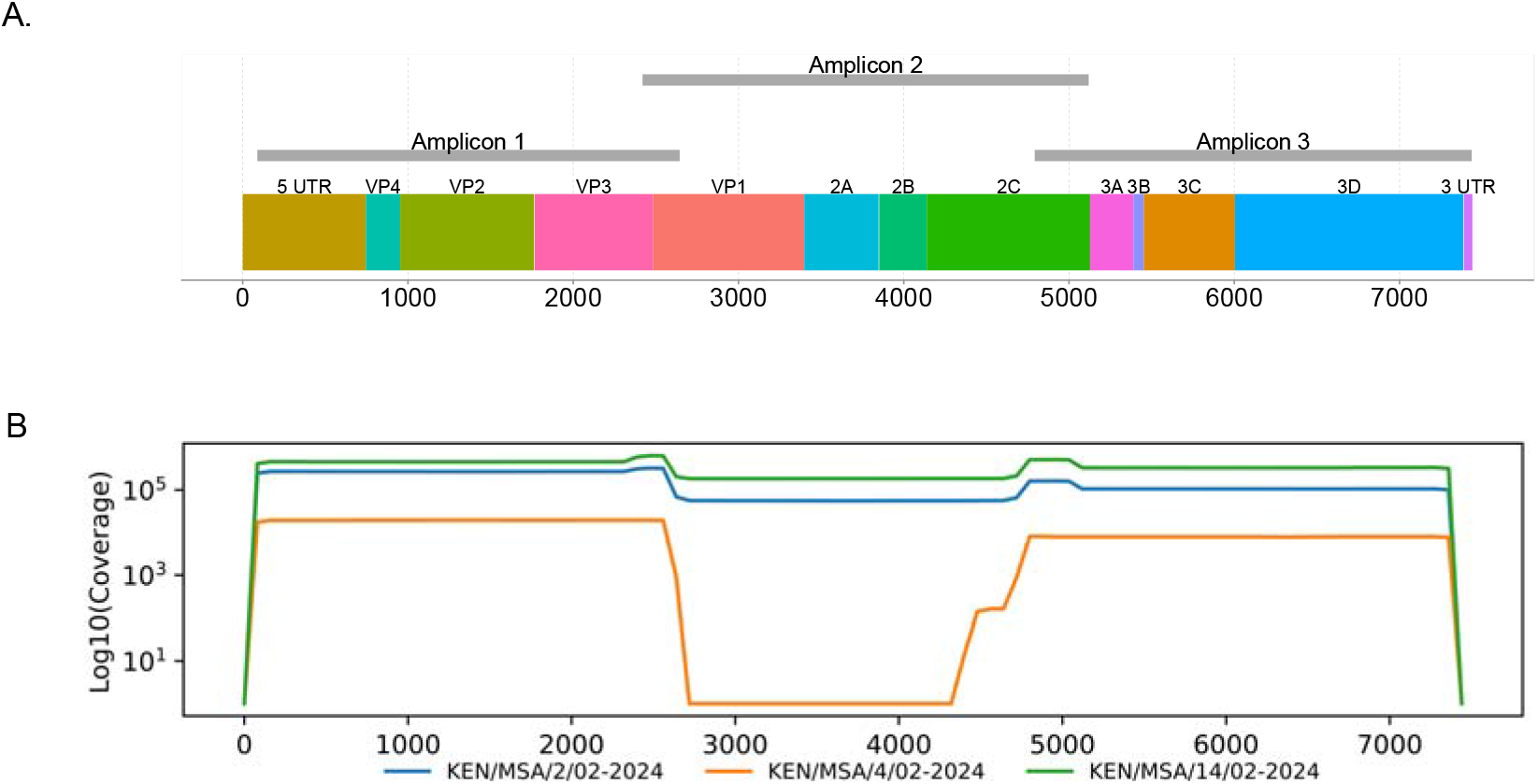
Genome maps. (A) Schematic representation showing the position of overlapping amplicons in the CA24v genome (GenBank accession number PP548240). (B) Coverage plots for KEN/MSA/2/02-2024, KEN/MSA/4/02-2024 and KEN/MSA/14/02-2024.

Viral RNA was extracted from the three ocular samples using the QIAamp Viral RNA Mini Kit (Qiagen) and reverse transcribed using the LunaScript® RT SuperMix Kit (New England Biolabs). A negative control (NC) (nuclease-free water) was included during both the extraction and reverse transcription steps. The cDNA was then amplified in three reaction tubes with the Q5® Hot Start High-Fidelity 2Master Mix (NEB) using the newly designed and optimised CA24v primers and thermocycling conditions as shown in **Table 1**. The PCR products were loaded on a 1.5% agarose gel to confirm amplification before purification using Agencourt AMPure XP beads. Library preparation was performed using the Ligation Sequencing Kit (SQK-LSK114) and Native Barcoding Kit (NBD96), and sequencing performed on the Oxford Nanopore Technologies (ONT) GridION platform.

Genome assembly was performed using a sub-workflow of an in-house pipeline named “*ViralPhyl*” and available on GitHub (https://github.com/kwtrp-peo/viralphyl). Base-called reads were demultiplexed using the ARTIC Guppyplex tool with default parameters, applying a minimum Q score of 9. Reads shorter than 500 nt were filtered out using the toullingQC module. Consensus sequences were generated by aligning the reads to a reference sequence (in this case CVA24_2400060741_FRA24, GenBank accession PP548240). The reads were aligned using MiniMap2 ^16^. Positions with genome coverage below 20 reads were masked with ‘N’. The resulting consensus sequences were further refined using Medaka v 2.0.1 to correct potential sequencing errors. The recovered genome sequences were combined with publicly available CA24 genomes and aligned using MAFFT v7.5201 ^14^. A maximum likelihood (ML) phylogenetic tree was inferred using IQ-TREE v2.1.3 (http://www.iqtree.org/) applying the GTR substitution model, with branch support assessed using 1000 bootstrap iterations. Nucleotide and amino acid variations between the newly sequenced genomes were analyzed using Snipit v1.6 ^17^. The analysis was performed with input options --sequence-type nt for nucleotide variation and --sequence-type aa for amino acid variation.

## Results and discussion

Two of the recovered genome sequences (KEN/MSA/2/02-2024 and KEN/MSA/14/02-2024) were 7,304 nucleotides (nt) in length **(Table 2)**, comprising the 5′ untranslated region (UTR) of 637 nt, complete open reading frame (ORF) of 6,645 nt, and 3′-UTR of 22 nt. Sequence KEN/MSA/4/02-2024 contained a 2,191- nucleotide gap within the ORF due to amplicon 2 dropout (Table 2, **Figure 1B**), which may have resulted from low viral load (Ct value > 38.43). Eight genotypes of CA24v are currently described (GI–GVIII). The new genomes generated here were classified as genotype IV, and clustered in clade comprising sequences sampled in France in February 2024 (**Figure 2A**). The recovered genomes displayed nucleotide variations (n=25) across the entire genome (**Figure 2B**). However, only synonymous mutations were observed (i.e., no amino acid substitutions) indicating conservation at the protein level despite genetic diversity.

**Table 2.**
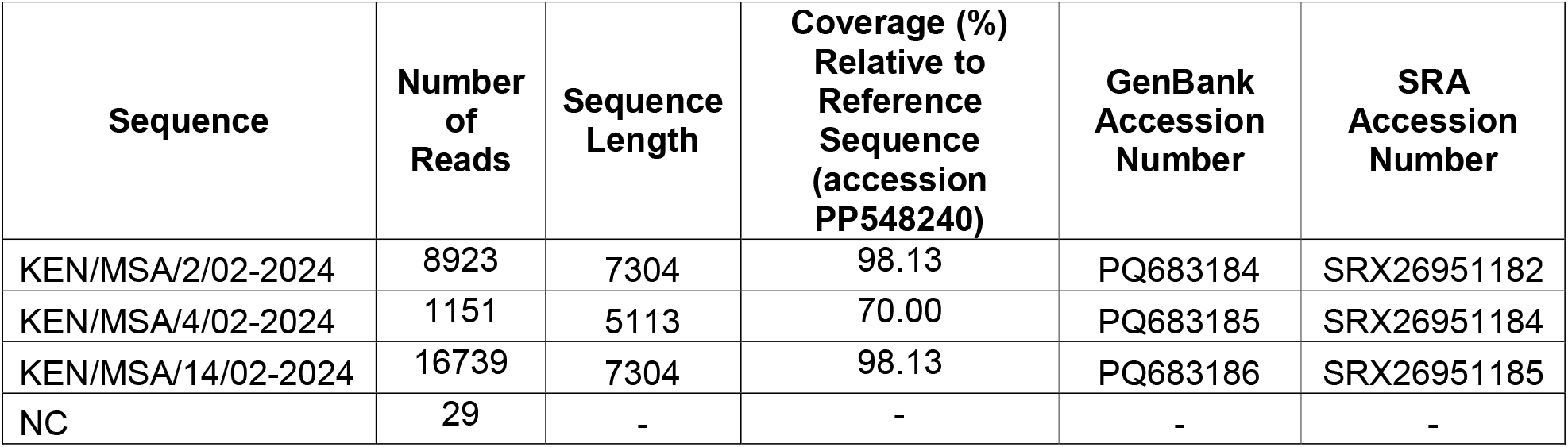
Genome length and coverage of the sequenced samples, along with their GenBank and SRA accession numbers.

**Figure 2.**
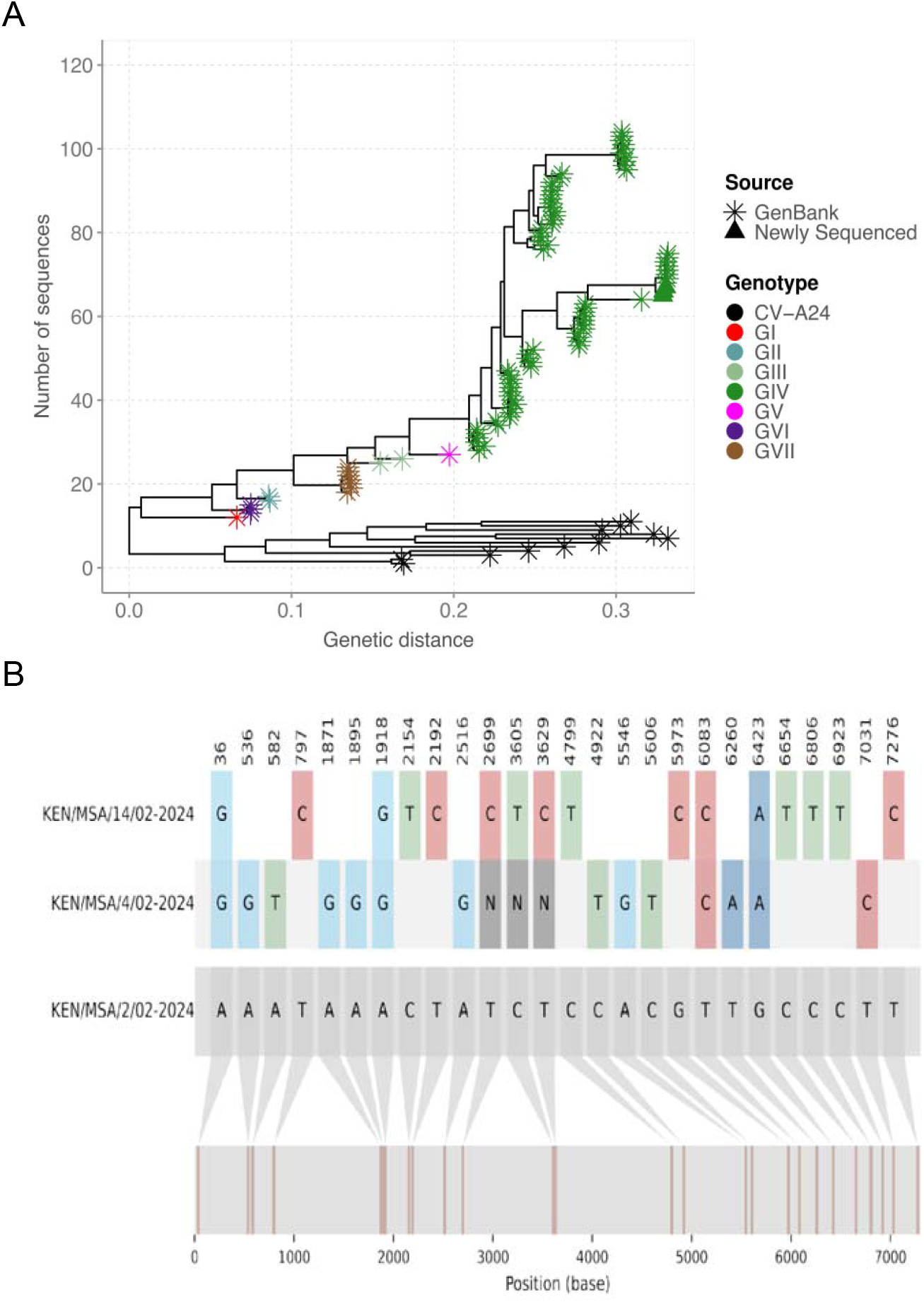
(A) Maximum likelihood phylogenetic tree based on the genome sequences of CA24v from this study (n=3) and previous outbreaks (n=119).(B) Nucleotide alignment showing the nucleotide variations across the three CA24v genomes, with KEN/MSA/2/02-2024 as the reference sequence.

This analysis protocol have some limitations. First, the selected primers did not capture the entire 5’ and 3’ UTR regions because the primers bind to regions within the UTRs, rather than at the terminal ends. Second, substantial genetic diversity exists within CA24v, yet our primers have only been tested with genotype IV which is the most commonly detected in recent studies. Future testing against a larger sample and diverse CA24v genotypes is needed to confirm similar performance across additional genotypes.

In summary, we describe a simple tiled-amplicon-based whole genome sequencing protocol for CA24 that has great potential to support future studies of CA24v genomic epidemiology.

## Acknowledgements

We are grateful to the individuals who provided the three clinical samples that were used to test the protocol and Ministry of Health for providing these samples to us. This working is published with permission from director KEMRI.

## Funding

Funding for this work is from a Wellcome (grant no. 226002/A/22/Z), The Rockefeller Foundation (Grant OXF-FDG01), and National Institute of Health and Care Research (grant. NIHR156467).

## Data availability

The genome sequences reported in this work are available in GenBank under accessions PQ683184 - PQ683186. The raw sequencing reads are available in NCBI’s Sequence Read Archive (SRA) under BioProject accession PRJNA1193512.

## Ethical approval

This study/analysis was reviewed and approved by KEMRI Scientific Ethics Review Unit (SERU) Committee based in Nairobi, Kenya (Protocol #: KEMRI/SERU/CGMR-C/304/4894).

## Competing interest statement

The authors have declared no competing interest.

## Notes

### Author Declarations

This study/analysis was reviewed and approved by KEMRI Scientific Ethics Review Unit (SERU) Committee based in Nairobi, Kenya (Protocol #: KEMRI/SERU/CGMR-C/304/4894).

## References

1. Shrestha R, RRK., KN., SO., RS., CR., SKL., & KS. Acute Conjunctivitis among Patients Visiting the Outpatient Department of Ophthalmology in a Tertiary Care Centre. Journal of Nepal Medical Association. 2024;62(269):24–26.

2. Tran H, Ha T, Hoang L, et al. Coxsackievirus A24 causing acute conjunctivitis in a 2023 outbreak in Vietnam. International Journal of Infectious Diseases. 2024;146. doi:10.1016/j.ijid.2024.107133

3. Shrestha E, Katuwal N, Kharel Sitaula R, et al. Investigation of the causative pathogen in the 2023 conjunctivitis outbreak of Nepal using unbiased metagenomic next generation sequencing. medRxiv. Published online January 1, 2024:2024.04.16.24305920. doi:10.1101/2024.04.16.24305920

4. Prajna NV, Prajna L, Teja V, et al. Apollo Rising: Acute Conjunctivitis Outbreak in India, 2022. Cornea Open. 2023;2(2). https://journals.lww.com/corneaopen/fulltext/2023/06000/apollo_risingacute_conjunctivitis_outbreak_in.1.aspx

5. Fonseca MC, Pupo-Meriño M, García-González LA, et al. Molecular evolution of coxsackievirus A24v in Cuba over 23-years, 1986–2009. Sci Rep. 2020;10(1):13761. doi:10.1038/s41598-020-70436-w

6. Haider SA, Jamal Z, Ammar M, Hakim R, Salman M, Umair M. Genomic Insights into the 2023 Outbreak of Acute Hemorrhagic Conjunctivitis in Pakistan: Identification of Coxsackievirus A24 Variant through Next Generation Sequencing. medRxiv. Published online January 1, 2023:2023.10.11.23296878. doi:10.1101/2023.10.11.23296878

7. Tran H, Ha T, Hoang L, et al. Coxsackievirus A24 causing acute conjunctivitis in a 2023 outbreak in Vietnam. International Journal of Infectious Diseases. 2024;146:107133. doi:10.1016/j.ijid.2024.107133

8. Yek C, Pacheco AR, Vanaerschot M, et al. Metagenomic pathogen sequencing in resource-scarce settings: Lessons learned and the road ahead. Frontiers in Epidemiology. 2022;2. https://www.frontiersin.org/journals/epidemiology/articles/10.3389/fepid.2022.926695

9. Quick J, Grubaugh ND, Pullan ST, et al. Multiplex PCR method for MinION and Illumina sequencing of Zika and other virus genomes directly from clinical samples. Nat Protoc. 2017;12(6):1261–1276. doi:10.1038/nprot.2017.066

10. Luka MM, Kamau E, de Laurent ZR, et al. Whole genome sequencing of two human rhinovirus A types (A101 and A15) detected in Kenya, 2016-2018. Wellcome Open Research 2021 6:178. 2021;6:178. doi:10.12688/wellcomeopenres.16911.1

11. Fall A, Abdullah O, Han L, et al. Enterovirus D68: Genomic and Clinical Comparison of 2 Seasons of Increased Viral Circulation and Discrepant Incidence of Acute Flaccid Myelitis—Maryland, USA. Open Forum Infect Dis. 2024;11(11):ofae656. doi:10.1093/ofid/ofae656

12. Zhang M, He D, Liu Y, et al. Complete genome analysis of echovirus 30 strains isolated from hand-foot-and-mouth disease in Yunnan province, China. Virol J. 2023;20(1):215. doi:10.1186/s12985-023-02179-9

13. Hu YF, Zhao R, Xue Y, Fan Y, Jin Q. Full Genome Sequence of a Novel Coxsackievirus B5 Strain Isolated from Neurological Hand, Foot, and Mouth Disease Patients in China. J Virol. 2012;86(20):11408–11409. doi:10.1128/jvi.01709-12

14. Katoh K, Standley DM. MAFFT multiple sequence alignment software version 7: improvements in performance and usability. Mol Biol Evol. 2013;30(4):772–780. doi:10.1093/molbev/mst010

15. Lambisia AW, Morobe JM, Moraa E, et al. Identification of coxsackievirus A24 variant during an acute hemorrhagic conjunctivitis outbreak in coastal Kenya, 2024. medRxiv. Published online January 1, 2024:2024.12.04.24318475. doi:10.1101/2024.12.04.24318475

16. Li H. Minimap2: pairwise alignment for nucleotide sequences. Bioinformatics. 2018;34(18):3094–3100. doi:10.1093/bioinformatics/bty191

17. O’Toole Á, Aziz A, Maloney D. Publication-ready single nucleotide polymorphism visualization with snipit. Bioinformatics. 2024;40(8):btae510. doi:10.1093/bioinformatics/btae510

